# Data processing pipelines and tools for routine health facility malaria surveillance in Uganda

**DOI:** 10.64898/2026.07.22.26357569

**Authors:** Austin Carter, Tom Eganyu, Dianna E.B. Hergott, John Rek, Jaffer Okiring, Daniel Kyabayinze, Catherine Maiteki, Paul Mbaka, David L. Smith

## Abstract

Timely processing of routine health facility surveillance data is essential for responsive malaria control, yet the path from raw electronic reports to actionable intelligence remains a largely undocumented challenge in endemic countries. We present an open-source Extract-Transform-Load (ETL) software system and accompanying metadata R package (`ramptools`) that together convert raw DHIS2 health facility data into cleaned, version-controlled, analysis-ready datasets for Uganda’s National Malaria Elimination Division (NMED). The ETL pipeline is implemented in R and deployed on a cloud platform with scripts running on automated schedules to keep the database current. The system extracts malaria indicators from two successive DHIS2 instances via the DHIS2 Web API, applies a two-stage outlier detection algorithm combining variance-based screening with STL decomposition, imputes missing values through seasonal interpolation, enforces logical consistency constraints across indicator cascades, and aggregates facility-level data through a six-level administrative hierarchy. All raw data are stored in an append-only versioned schema, enabling reconstruction of the database state at any historical point. The `ramptools` package provides standardized metadata—including an indicator crosswalk mapping 118 indicators across DHIS2 instances, a location hierarchy of 11,229 organizational units, geolocated health facility attributes, and administrative boundary shapefiles—as lazy-loaded R data objects. We validate the system through a proof-of-principle outbreak detection application that consumes ETL outputs to compute district-level outbreak indices via kernel-smoothed time series analysis, deployed as an interactive Shiny dashboard. The software has been in development since 2020, processing weekly and monthly data for approximately 8,700 health facilities. All code is open source (MIT license) and hosted on GitHub.

## Introduction

### The data-to-decision gap in malaria control

Routine surveillance data are collected to support responsive public health policy, but the practical infrastructure for timely, evidence-based review of information, particularly in lower-income countries where disease burden is highest, remains underdeveloped. For malaria, the dominant modality of resource allocation has recently operated on multi-year cycles: three-year disbursement periods for Global Fund support [1], annual estimates in the World Malaria Report [2], and five-year strategic plan revisions. These cycles involve collation of available data by external contractors, calibration of transmission models, and submission of funding requests months or years behind the data that entered the analyses. This lag, and the reliance on external expertise, limits application to long-term strategic planning and prevents responsive management—for example, detecting and responding to outbreaks within weeks of their onset [3].

### Electronic reporting and unmet potential

The introduction of electronic health management information systems (HMIS), particularly the open-source DHIS2 platform [4, 5], created the technical possibility of near-real-time access to facility reporting data. As of 2024, DHIS2 is deployed in over 100 countries and serves as the primary health information system in much of sub-Saharan Africa [6]. Uganda introduced its first DHIS2 instance in 2013, with a second instance deployed in 2020 incorporating revised indicator definitions and reporting forms. However, the existence of an electronic data platform does not, by itself, solve the analytical challenges that stand between raw reports and usable intelligence. Health facility data suffer from well-documented problems including incomplete reporting, data entry errors arising from translating paper records into an electronic record, logical inconsistencies between related indicators, and changes to indicator definitions across system migrations [7–10]. These problems are compounded when analysts must independently access DHIS2, perform ad hoc cleaning, and reconcile data across system versions—leading to methodological inconsistencies across reports and duplicated effort (Box 1) [11].

##### Standardizing the data processing pipeline

Without a standardized data processing pipeline, every project would need to develop its own method for data extraction and data cleaning: every analyst would extract some set of malaria indicators from DHIS2, identify outliers, run consistency checks, and impute values for missing or outliered data before conducting an analysis. Documentation and archiving were also the responsibility of individual analysts. Differences in the outputs of analyses could be due to differences in data cleaning or to the timing of the extraction.

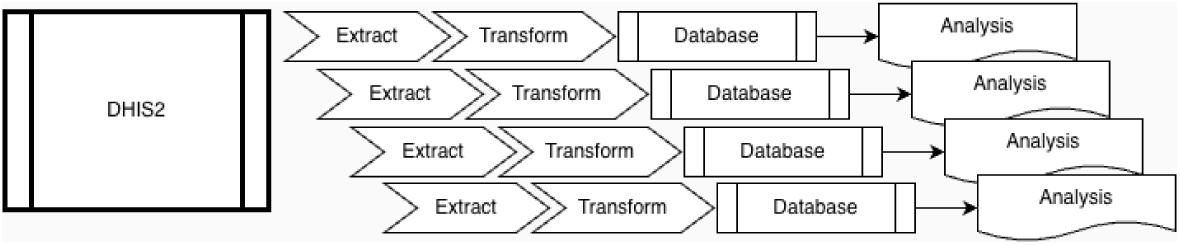

After standardizing the data processing pipelines, raw data are extracted from DHIS2 and stored in an append-only database, so it is fully auditable. The production algorithms that transform the data are open source, so users can examine the raw data and contribute to development of improved algorithms. All users have access to the clean malaria indicators, eliminating heterogeneity in studies that is caused by heterogeneity in the handling of the data.

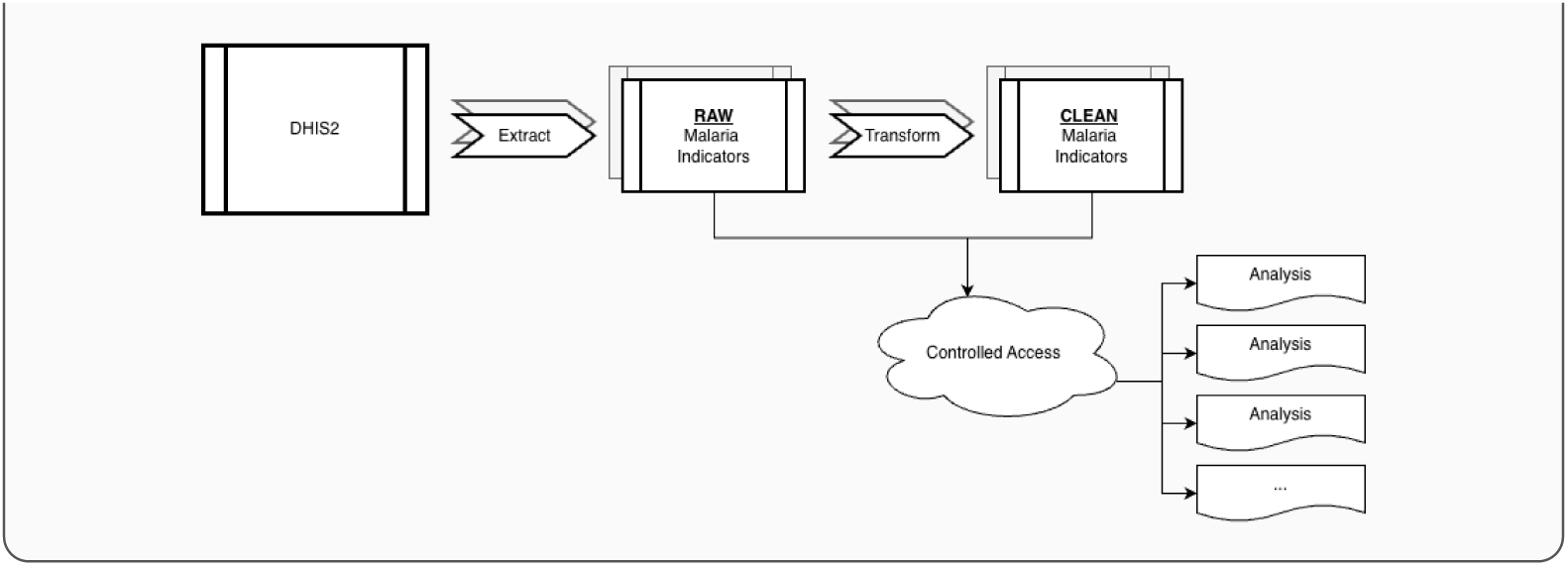

### Prior work on health facility data processing

Several efforts have previously addressed aspects of health facility data quality and processing. The WHO Data Quality Review (DQR) toolkit provides a framework for periodic assessment of data quality dimensions including completeness, internal consistency, and external consistency to support WHO-related analysis [12]. Routine Data Quality Assessment (RDQA) tools have been developed for facility-level auditing [13]. Automated approaches to outlier detection in DHIS2 data have been explored using statistical methods including z-scores, modified z-scores, and interquartile range methods [14]. Imputation methods for missing health facility data have been examined, with approaches ranging from simple carry-forward to model-based spatial and temporal interpolation [15, 16]. The `dhis2R` and `datimutils` R packages provide programmatic access to DHIS2 APIs [17], and several country programs have developed internal data processing workflows [18]. The Malaria Atlas Project and the Institute for Health Metrics and Evaluation have developed pipelines for incorporating facility data into geospatial models, though these serve estimation rather than routine program management [19, 20].

What has been largely absent from the literature is a comprehensive, openly available practical system that integrates extraction, cleaning, versioning, metadata management, and deployment into a single framework designed for routine use by a national malaria program. Existing tools address individual components—API access, outlier detection, or spatial analysis—but the end-to-end pipeline connecting raw DHIS2 data to analyst-ready datasets, with full provenance tracking and automated scheduling, has not been described or made available as reusable software.

### The RAMP Uganda collaboration

We thus developed open source software to create an Extract-Transform-Load data processing pipeline that creates stable up-to-date malaria data for Uganda’s Ministry of Health. The project was a collaboration between the University of Washington, Uganda’s Department of Health Information (DHI), and the National Malaria Elimination Division (NMED) as part of the Robust Analytics for Malaria Policy (RAMP) program. The collaboration was prompted by three practical needs: (1) reducing demand on DHIS2 servers from repeated analyst queries, (2) standardizing data processing to eliminate inconsistencies across reports, (3) enabling timely analysis by providing cleaned data shortly after each reporting deadline; and (4) providing analysts with a set of computing resources and utilities. The resulting system comprises two open-source R packages (`uga-etl-facility-data` and `ramptools`), a cloud deployment infrastructure, and a downstream outbreak detection application that serves as both a validation of the pipeline outputs and an operational tool for the program.

This manuscript presents the design, implementation, and validation of this system. We describe the software architecture and algorithms, the metadata standards and database schema, the deployment and maintenance framework, and a proof-of-principle outbreak detection application. All code is open source under the MIT license and available on GitHub (https://github.com/dd-harp).

## Design and implementation

### System architecture

The system consists of three components (Fig 1):

1. `ramptools` (R package, v0.2.0): Provides standardized metadata tables, spatial data objects, and database access functions. Available at https://github.com/dd-harp/ramptools.
2. `uga-etl-facility-data` (R scripts + cloud deployment): Implements the ETL pipeline—extraction from DHIS2, data cleaning and transformation, and loading into the data warehouse. Available at https://github.com/dd-harp/uga-etl-facility-data.
3. `outbreak` (R scripts + Shiny application): A downstream application that consumes cleaned data to compute and visualize district-level malaria outbreak indices. Available at https://github.com/dd-harp/outbreak.

**Fig 1.**
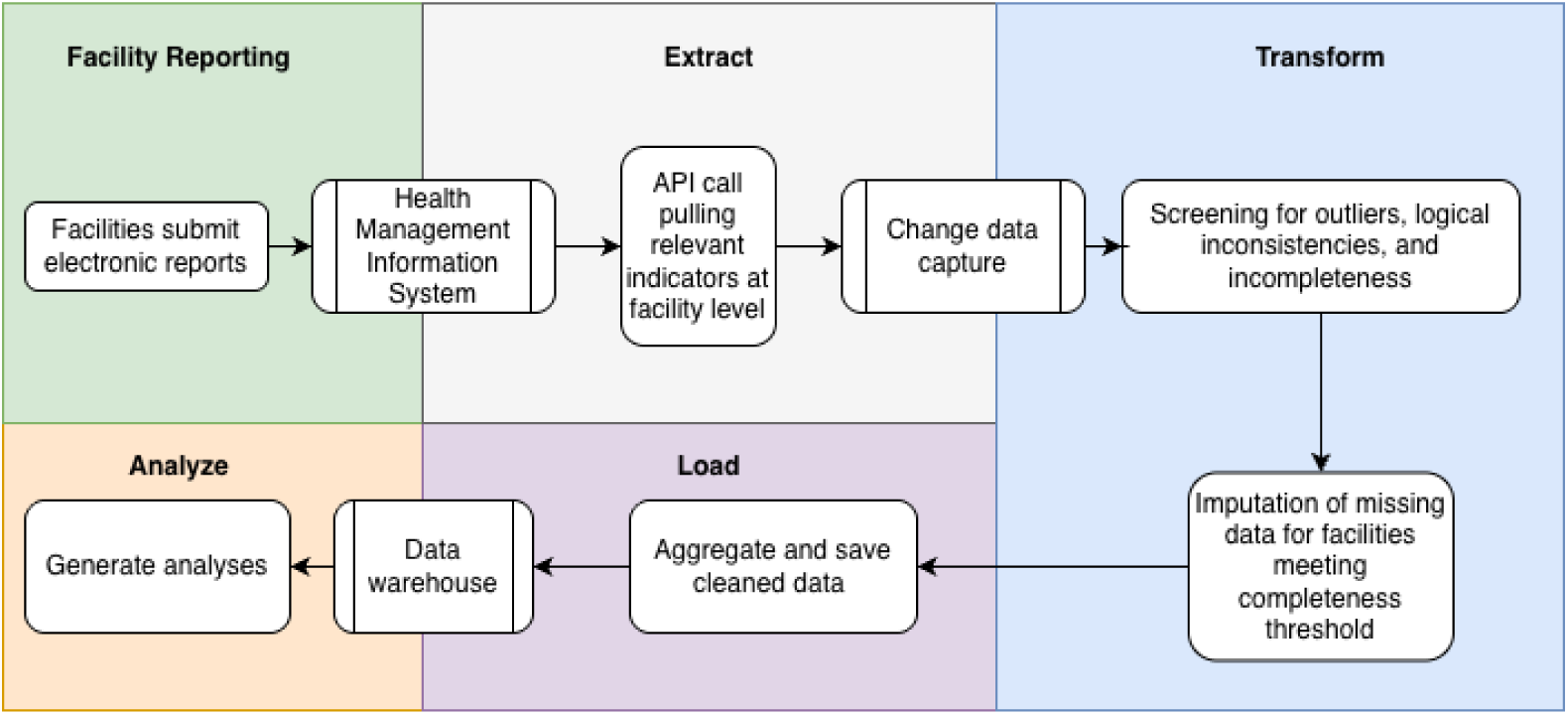
System architecture of the Uganda malaria facility data ETL pipeline. Data flows from two DHIS2 API instances through the ETL pipeline (extraction, change data capture, outlier detection, imputation, consistency checks, aggregation) into versioned storage in a data warehouse. Downstream applications (outbreak dashboard, routine reports) consume cleaned data from from the data warehouse. Rectangles represent data stores, rounded rectangles represent processes, and parallelograms represent outputs.

The system is implemented entirely in R (version ≥ 3.5.0) with dependencies managed via `renv` [21]. Key R package dependencies include `data.table` for data manipulation, `httr` and `jsonlite` for API interaction, `forecast` for time series methods [22], `bigrquery` for cloud database access, and `sf` for spatial operations. The cloud deployment uses Docker containers on Google Cloud Platform.

### Data sources

Uganda operates two successive DHIS2 instances: Instance 1 (2013–2019), with indicator identifiers specific to the original deployment, and Instance 2 (2020–present), with revised indicator definitions, identifiers, and reporting forms. The ETL system unifies data from both instances through a crosswalk table that maps instance-specific identifiers to canonical indicator codes, enabling seamless time series analysis across the transition. Four reporting forms contain malaria-relevant indicators (Table 1).

**Table 1.** DHIS2 reporting forms processed by the ETL.

| Form | Frequency | Description |
| --- | --- | --- |
| 033b | Weekly | SMS-submitted subset of key indicators |
| 105 | Monthly | Outpatient department, all indicators |
| 108 | Monthly | Inpatient department, all indicators |
| 097b | Quarterly | Facility-aggregated village health team (VHT) data |

### The ramptools metadata package

A central challenge in working with DHIS2 data is that data dimensions—who, what, when, and where—are encoded in DHIS2-specific formats that require translation for analysis. The `ramptools` R package provides this translation layer through curated metadata tables and utility functions, distributed as lazy-loaded R data objects for zero-configuration access.

#### Indicator table

The indicator table (118 rows × 6 columns) maps every malaria-relevant DHIS2 data element to a canonical representation (Table 2). Each row contains: the DHIS2 data element name, a human-readable label, a short code for programmatic use (`code name`), the DHIS2 unique identifier, the reporting frequency, and the DHIS2 instance version (1 or 2). The same canonical `code name` (e.g., `rdt pos`) maps to different identifiers across instances, enabling the pipeline to query either instance using a single set of indicator references. Indicators span categories including outpatient attendance, malaria case counts (suspected, confirmed, severe), diagnostic testing (RDT and microscopy), treatment, commodity stock status, antenatal care and intermittent preventive treatment, inpatient admissions, and community health worker reports. The complete indicator table is provided in **??**.

**Table 2.** Structure of the indicator crosswalk table. Example entries showing how a single canonical code name maps to different DHIS2 identifiers across instances.

| code_name | display_name | dhis_id (v1) | dhis_id (v2) | frequency |
| --- | --- | --- | --- | --- |
| rdt_pos | RDT Positive | KPmTI3TGwZw.PfdPCxOoiBj | OOsYcFK5YbS | monthly |
| conf_malaria | Confirmed Malaria | [v1 ID] | [v2 ID] | monthly |
| al_stockout | AL Stock-out | [v1 ID] | [v2 ID] | monthly |

#### Location hierarchy

The location table (11,229 rows × 9 columns) encodes Uganda’s complete administrative and health facility hierarchy across six levels: national (1 unit), region (15 units), district (∼146 units), district local government (DLG), subcounty (∼2,204 units), and health facility (∼8,676 units). Each entry records the organizational unit name, DHIS2 identifier, parent identifier, level, and the full path to the root. Pre-computed columns provide the region, district, DLG, and subcounty names for each entity, enabling efficient hierarchical queries without recursive traversal.

#### Health facility attributes

The health facility table (8,676 rows × 22 columns) extends the location hierarchy with facility-specific attributes: facility level (Health Centre II/III/IV, Hospital, Clinic), ownership (government, private for-profit, private not-for-profit), medical bureau affiliation, operational and reporting status, and geographic coordinates (latitude, longitude). A `geom enhanced` flag identifies facilities whose coordinates were obtained through supplementary geocoding rather than from DHIS2 metadata.

#### Demographic disaggregation

The age-sex table (14 rows × 4 columns) maps DHIS2 category option combo identifiers to structured demographic fields (sex: male/female; age groups: 0–4, 5–9, 10–19, 20+, and neonatal categories), enabling extraction at the most granular available level.

#### Spatial data

Four `sf` spatial data objects provide administrative boundary geometries: district-level polygons (∼146 features), subcounty-level polygons (2,204 features with quality flags), region-level polygons (15 features), and water body geometries for cartographic overlays. These shapefiles maintain topological consistency: child geometries tile their parent without gaps or overlaps, each child belongs to exactly one parent, all geometries are topologically valid, and parent territories are fully covered with remainder polygons where needed.

#### Period utilities

Functions `make week map()` and `make month map()` convert DHIS2 period strings (e.g., `“2020W17“`, `“202004“`) to structured date representations with start, end, and midpoint dates, handling ISO week edge cases at year boundaries. The function `get period range()` generates DHIS2-formatted period vectors for API queries.

#### Database access functions

The package provides parallel sets of functions for two storage backends. The BigQuery production backend includes: `bq connect()`, `bq get data()`, `bq get clean data()`, `bq get db diff()`, `bq get latest version()`, `bq append raw data()`, `bq write clean data()`, `bq write imputed data()`, and `bq init tables()`. A SQLite legacy backend provides equivalent functions for local development. Both backends implement the same versioned, append-only storage pattern.

### Extract: data acquisition from DHIS2

#### API interaction

Data extraction is performed through the DHIS2 Analytics Web API endpoint (`/analytics/dataValueSet.json`). The `pull data()` function orchestrates the extraction: (1) DHIS2 credentials are loaded from a JSON configuration file (or environment variables in cloud deployment) and validated via a test API call; (2) API query URLs are assembled with three dimensions—data elements, time periods, and organizational units—drawn from the metadata tables; (3) organizational units are requested in batches of 50 to respect API payload limits; (4) administrative metadata columns are removed, retaining only dimensions and values needed for analysis.

#### Change data capture

Rather than replacing the entire database on each extraction, the system implements change data capture (CDC). The `bq get db diff()` function performs an anti-join between freshly extracted data and the current database contents, returning only rows that are new or have changed values. This differential is appended to the raw data table with a monotonically increasing version number, preserving the complete history of observations.

#### Extraction schedule

Extractions are scheduled to align with reporting deadlines: weekly data is extracted every Tuesday at 06:00 UTC (most recent 5 weeks), monthly data on the 5th of each month at 06:00 UTC (most recent 12 months). The overlapping extraction windows ensure that late submissions and retroactive corrections are captured through the CDC mechanism. Each extraction records provenance metadata: version number, timestamp, user identity, git commit hash, git branch, and the time period range covered.

### Transform: data cleaning and quality assurance

The transformation stage applies four sequential operations: completeness assessment, outlier detection, imputation, and logical consistency enforcement.

#### Completeness assessment

The `completeness()` function evaluates reporting completeness at the facility-indicator level—a finer granularity than the facility-level reporting rate available in DHIS2. For each facility-indicator pair, the function expands observed data to a complete grid of all expected time periods between the facility’s first and last report for that indicator, then computes the completeness ratio:

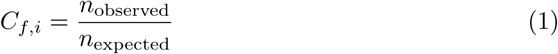

where *n*_expected_ spans from first to last report. Facility-indicator pairs with *C_f,i_ <* 0.5 are excluded from imputation to avoid interpolating across excessively sparse data. This approach distinguishes three types of missingness (Fig 2): (1) periods before a facility’s first report (structural), (2) gaps between the first and last report (potentially correctable), and (3) periods after the last report (the facility may have ceased operations).

**Fig 2.**
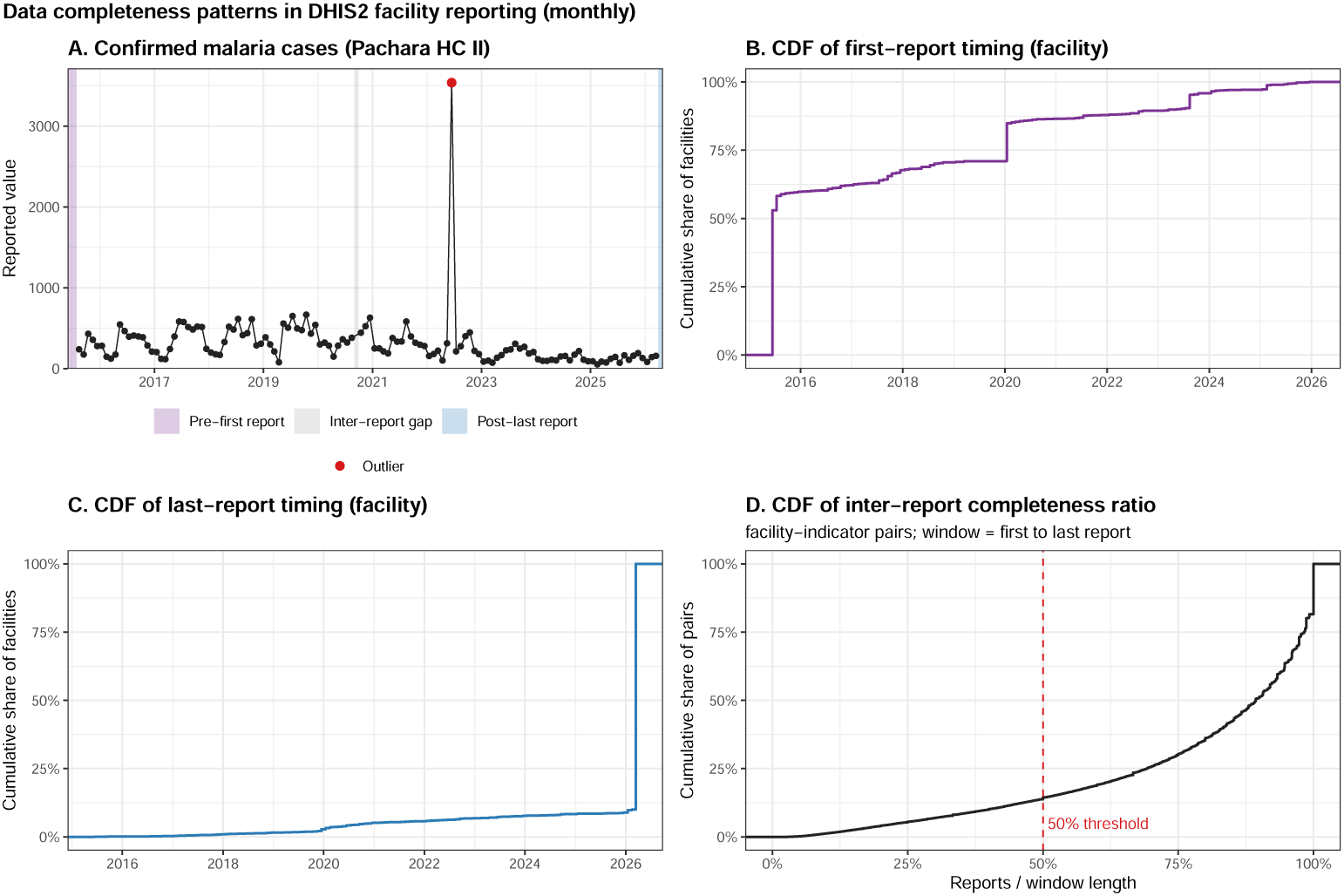
Data completeness patterns in health facility reporting. A: Example facility time series showing three types of missingness—pre-first-report (purple), inter-report gaps (grey), and post-last-report (blue)—with outliers highlighted in red. B: Cumulative distribution of timing of first report across facilities. C: Cumulative distribution of timing of last report. D: Cumulative distribution of inter-report completeness ratio across facility-indicator pairs, with vertical line at the 50% exclusion threshold.

#### Outlier detection

The `outlier and impute()` function implements a two-stage outlier detection algorithm applied independently to each facility-indicator time series:

##### Stage 1—Variance-based screening

Values satisfying |*x_t_* − *x̃*| *>* 5 · *s_x_* are flagged, where *x̃* is the median and *s_x_* is the standard deviation of the series. This captures extreme entry errors (e.g., an extra zero) that would distort subsequent seasonal decomposition.

##### Stage 2—Seasonal decomposition

The `tsclean()` function from the R `forecast` package [14] applies STL (Seasonal and Trend decomposition using Loess) [23] to decompose the series into trend, seasonal, and remainder components. Observations whose remainder component exceeds three times the interquartile range above the 75th percentile (or below the 25th percentile minus the IQR) are flagged as outliers. For series too short for seasonal decomposition, Friedman’s super smoother is used instead.

Flagged values are set to `NA` and treated as missing for imputation. All outlier flags are recorded in the output, preserving the original values alongside imputed replacements.

#### Imputation

Missing values (including those created by outlier removal) are imputed using the seasonal interpolation method implemented in `forecast::tsclean()`. This applies linear interpolation to the seasonally-adjusted series, then re-adds the seasonal component:

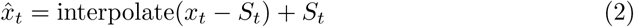

where *S_t_* is the estimated seasonal component from STL decomposition. Imputed values are floored at zero to maintain epidemiological plausibility. Imputation is restricted to the period after each facility’s first report, and only for facility-indicator pairs meeting the 50% completeness threshold. The pipeline retains both original and imputed values, enabling analysts to assess the impact of imputation on any analysis.

#### Logical consistency enforcement

The `inconsistency()` function checks relationships between indicators that must hold by definition. These relationships are encoded in a configuration table specifying parent-child indicator pairs where the child value must not exceed the parent (Table 3). Violations are flagged for review. In the imputation stage, cascade relationships are respected by modeling parent indicators as counts and child indicators as proportions of their parents.

**Table 3.**
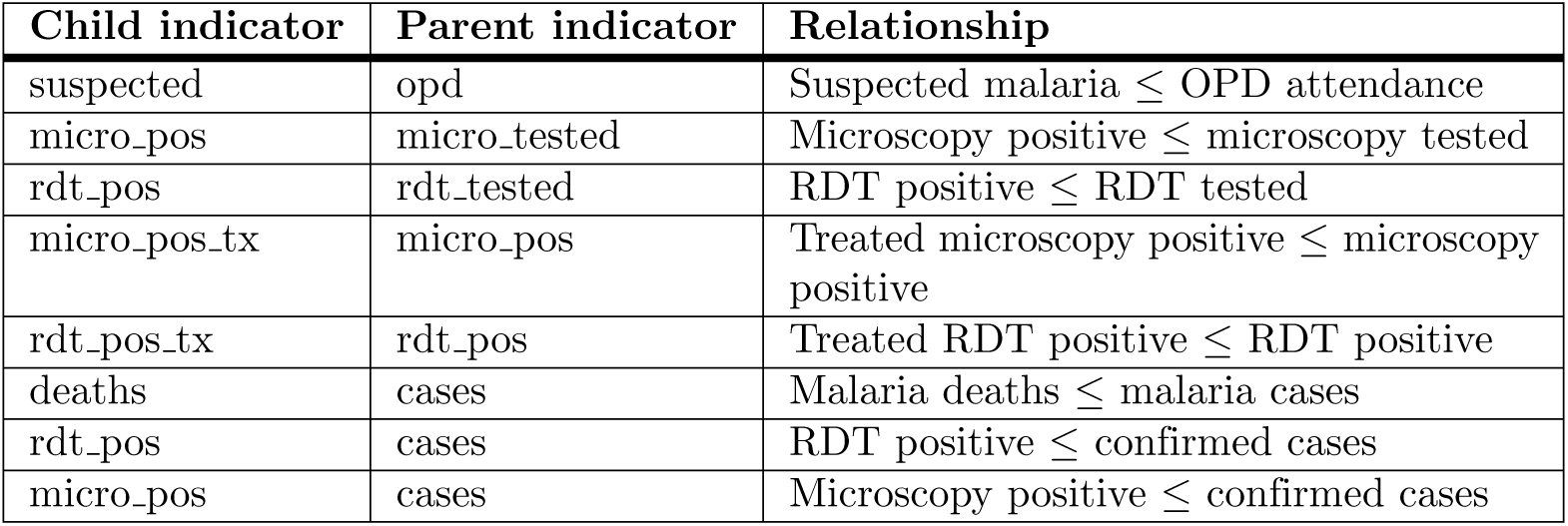
Logical consistency constraints enforced by the ETL. Each row defines a parent-child relationship where the child indicator must not exceed the parent.

### Aggregation

The `agg to parent()` function aggregates facility-level data upward through the administrative hierarchy by summing both original and imputed values at each level. Aggregation proceeds iteratively through five steps: facility → subcounty → DLG → district → region → national. The function merges with the location table to obtain parent identifiers at each level, enabling automated aggregation that adapts to changes in the administrative hierarchy without code modifications.

### Load: data warehouse and versioning

#### Database schema

The production data warehouse uses Google BigQuery (project: `uganda-malaria`, dataset: `uga facility data`). For each reporting frequency (weekly, monthly), four tables are maintained (Table 4).

**Table 4.** BigQuery data warehouse schema. Four tables per reporting frequency support versioned raw storage and current cleaned outputs.

| Table | Columns | Write mode | Purpose |
| --- | --- | --- | --- |
| `raw_{freq}_data` | `dataElement`, `period`, `orgUnit`, `value`, `timestamp`, `version` | Append-only | Complete history of extracted values |
| `raw_{freq}_version_metadata` | `version`, `timestamp`, `user`, `commit`, `branch`, `period_start`, `period_end`, `api_call` | Append-only | Extraction provenance |
| `clean_{freq}_data` | `location_id`, `location_name`, `period`, `code_name`, `value`, `imputed_value`, `level` | Overwrite | Current cleaned, aggregated data |
| `imputed_{freq}_facility_data` | `location_id`, `date_mid`, `period`, `value`, `imputed_value`, `is_outlier`, `level`, `code_name` | Overwrite | Facility-level imputation details |

The raw tables use an append-only pattern: data is never overwritten or deleted. Each observation carries a version number, and the current state is reconstructed through window functions that select the most recent version of each unique observation (`ROW NUMBER() OVER (PARTITION BY … ORDER BY version DESC)`). This design enables reconstruction of the database as it appeared at any historical version.

The clean tables are overwritten on each processing run, reflecting the latest application of the cleaning algorithms to the complete raw data history. This separation ensures that raw data integrity is never compromised by changes to cleaning logic, while analysts always access the most current cleaned data.

A SQLite backend is maintained for local development and environments without cloud access. The `diff.R` module provides functions for creating, applying, and reversing data diffs—enabling forward and backward reconstruction of any historical database state from a sequence of incremental changes.

### Cloud deployment

#### Container and infrastructure

The ETL is deployed as a Docker container (base image: `rocker/r-ver:4.3.2`) with all system dependencies (libcurl, libssl, libxml2, libgit2, GDAL/GEOS/PROJ for spatial operations) and R packages pre-installed. The container installs `ramptools` directly from GitHub, ensuring the deployed version matches the latest release.

#### Scheduling and execution

Two Google Cloud Run Jobs execute the pipeline on schedule. The weekly job runs every Tuesday (cron: `0 6 * * 2`), extracting and cleaning weekly data. The monthly job runs on the 5th of each month (cron: `0 6 5 * *`), extracting and cleaning monthly data. Both are configured with 4 GB RAM, 2 CPUs, and a 1-hour timeout. Cloud Scheduler triggers these jobs automatically. DHIS2 credentials are stored in Google Secret Manager and injected as environment variables at runtime, eliminating credential storage in code or containers.

#### Cost and access

The cloud infrastructure operates at minimal cost (estimated *<*$5/month for BigQuery storage and Cloud Run execution). A dedicated service account (`dhis-etl-runner`) holds only the minimum required IAM roles (BigQuery Data Editor, Secret Manager Secret Accessor). Data access is controlled through BigQuery dataset-level permissions.

### Diagnostic visualization

The `plot facility data()` function generates multi-page PDF diagnostic reports for each district. For each district, the report includes: (1) a summary page listing the count of outliers detected and missing values imputed; (2) an aggregate district-level time series showing the combined cleaned signal; (3) paginated facet plots (3×4 or 4×4 grids) for each facility, showing raw values (black), imputed values (blue), outlier flags (red), and missing periods (grey shading). These reports enable domain experts to visually validate algorithmic decisions. An example is provided in **??**.

## Validation: outbreak detection application

Access to clean, up-to-date data benefits a malaria program, but it is an essential component of developing a system for outbreak identification and response. To demonstrate that the ETL pipeline produces data suitable for operational analytics, we present a proof-of-principle outbreak detection system that depends on cleaned ETL outputs and that outputs a timely oubtreak report, currently used by Uganda’s NMED.

### Outbreak index computation

The outbreak detection algorithm operates on district-level monthly confirmed malaria case counts retrieved from BigQuery via `bq get clean data()`. For each district-indicator time series, the algorithm:

1. Computes kernel-smoothed estimates at four bandwidths using Gaussian kernel smoothing (`ksmooth()`): *b*_60_ (60-day, short-term trend), *b*_100_ (100-day, medium-term trend), *b*_365_ (365-day, long-term trend), and *b*_0_ (median, overall baseline).
2. Estimates seasonal adjustment: for each calendar month, computes the median ratio of medium-term to long-term trends (*b*_100_*/b*_365_) across all years, yielding a relative seasonality factor.
3. Computes the outbreak index:

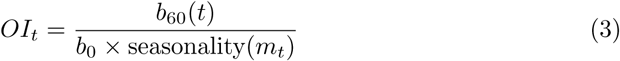

where *m_t_* is the calendar month of time *t*. Districts with *OI_t_ >* 2.0 (default threshold, user-adjustable) are flagged. This approach is compared against a WHO-style reference method that flags districts where the current month’s value exceeds the 75th percentile of historical values for the same calendar month.

### Interactive dashboard

The outbreak detection system is deployed as a Shiny web application (https://aucarter.shinyapps.io/outbreak_detection/) providing: an interactive choropleth map of Uganda districts colored by outbreak index on a log scale; a date slider with animation for temporal navigation (monthly steps from 2018 onward); an adjustable outbreak threshold (1.0–5.0); selection of indicator (confirmed malaria cases, malaria admissions, or a combined index); and a ranked table of districts exceeding the current threshold (Fig 3).

**Fig 3.**
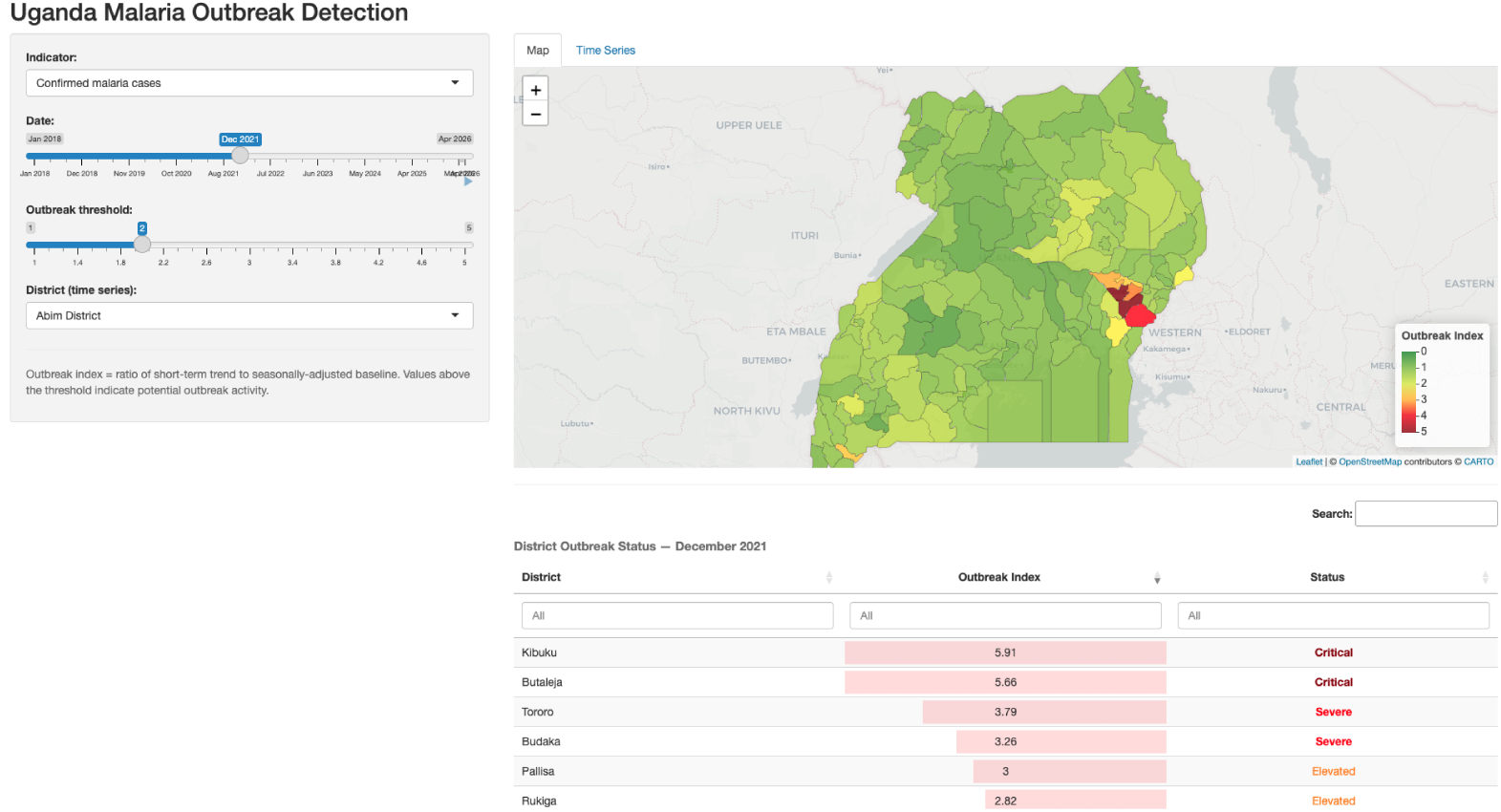
Outbreak detection Shiny dashboard. Screenshot of the interactive web application showing a choropleth map of Uganda districts colored by outbreak index (green–yellow–red gradient on a log scale). Controls include a date slider for temporal navigation, threshold adjustment, and indicator selection. A table of districts exceeding the threshold is displayed alongside the map.

The dashboard loads data directly from BigQuery on startup (with a pre-computed fallback dataset for resilience), demonstrating end-to-end integration from DHIS2 extraction through cleaning to analyst-facing visualization.

## Availability and maintenance

This ETL system was designed to provide timely responses while retaining the ability to adapt to malaria programmatic needs. The source code thus clearly defines the current operating version, but all users have access to the algorithms, to the raw data (the aligned indicators from the two instances of DHIS2), and to the cleaned data. Any modifications to the current operating version undergo extensive testing to ensure interoperability and continuous up time. All analysts are encouraged to follow the same practices to ensure replicability and hold analysts accountable for the advice they provide to the Ministry of Health. The following describes the protocols and procedures that enforce these principles.

### Source code

All source code is openly available under the MIT license:

- *ramptools*: https://github.com/dd-harp/ramptools
- *uga-etl-facility-data*: https://github.com/dd-harp/uga-etl-facility-data
- *outbreak*: https://github.com/dd-harp/outbreak

The repositories use `renv` for R dependency management, ensuring reproducible environments.

### Dependencies

The software requires R ≥ 3.5.0 and depends on the following open-source R packages: `data.table`, `httr`, `jsonlite`, `forecast`, `ggplot2`, `ggforce`, `sf`, `bigrquery`, `gargle`, `DBI`, `RSQLite`, `dplyr`, `lubridate`, `yaml`, and `devtools`. Cloud deployment requires Docker and access to Google Cloud Platform services (Cloud Run, BigQuery, Secret Manager, Cloud Scheduler, Artifact Registry). The SQLite backend provides a fully functional local alternative for environments without cloud access.

### Installation and usage

To install `ramptools`:

`devtools :: install_github (“dd - harp / ramptools”)`

To run the ETL locally, users clone the `uga-etl-facility-data` repository, configure DHIS2 credentials in a `dhis2 credentials.json` file, and execute the appropriate script (e.g., `scripts/clean aggregate monthly.R`). For cloud deployment, the `cloud/deploy.sh` script automates the complete GCP setup: creating the Artifact Registry, building and pushing the Docker image, configuring the service account with appropriate IAM roles, creating Cloud Run Jobs, and scheduling execution via Cloud Scheduler.

### Long-term maintenance

Technical stewardship of the codebase is shared between the University of Washington and Uganda’s Department of Health Information (DHI). The maintenance framework includes:

- *Version control.* All algorithms are stored in GitHub repositories. Changes to ETL logic are tracked through git commits, and the version metadata table records the exact commit hash used for each data extraction, ensuring full reproducibility.
- *Algorithm review cycle.* The cleaning algorithms undergo periodic review by both technical and domain teams. Changes are proposed through GitHub issues, implemented on feature branches, and merged after review—enabling the pipeline to evolve while maintaining an auditable history.
- *Metadata updates.* The location hierarchy and indicator tables are updated when new facilities are added, administrative regions change, or indicator definitions are revised. Source CSVs in `data-raw/` are regenerated and packaged into new releases.
- *Infrastructure hosting.* Cloud infrastructure (BigQuery storage, Cloud Run compute, Secret Manager) is hosted in the `uganda-malaria` GCP project. DHI maintains administrative access, ensuring continuity independent of external collaborators.
- *Community contributions.* Bug reports and feature requests are managed through GitHub Issues. The MIT license permits unrestricted reuse and adaptation by other country programs.

## Discussion

### Contributions and novelty

The system described here addresses a gap between the availability of electronic health facility data in DHIS2 and its usability for routine malaria program management. While individual components of our pipeline—API access, outlier detection, imputation, spatial analysis—have precedents, the integration of these components into a single, automated, version-controlled framework designed for production use by a national malaria program represents a novel contribution. Several features distinguish this work.

First, the unified indicator crosswalk addresses a common challenge: migration between DHIS2 instances typically creates a discontinuity in time series that requires manual reconciliation by each analyst. Our indicator table provides a single, maintained mapping enabling seamless analysis across Uganda’s 2013–present data history.

Second, the append-only versioned storage with provenance tracking provides full auditability of how the database has changed over time. This is critical for reproducing past analyses and understanding how policy recommendations may have evolved as data was corrected or resubmitted.

Third, the architectural separation between immutable raw storage and regenerable clean outputs ensures that improvements to cleaning algorithms can be applied retroactively to the complete data history without risking data loss.

Fourth, the containerized Cloud Run deployment with scheduled execution eliminates the need for manual pipeline execution and reduces the lag between reporting deadlines and data availability. The minimal cost (*<*$5/month) makes this infrastructure accessible to resource-constrained programs.

### Comparison with existing approaches

The WHO Data Quality Review toolkit [12] provides a comprehensive framework for periodic data assessment but is designed for manual, retrospective evaluation rather than automated, routine processing. Our system complements the DQR by implementing automated versions of several DQR checks as part of a continuous pipeline. The `dhis2R` package [17] provides DHIS2 API access but not the downstream cleaning, versioning, or deployment infrastructure. Country-specific data processing workflows exist but are typically not published, not open source, or not designed for reuse [18]. Alegana et al. [15] discuss imputation methods for missing facility data, from facility-level moving averages to Bayesian spatial-temporal models, while noting these address research-grade burden estimation rather than the underlying data-quality problems facing national malaria programs.

### Limitations

The current system has several limitations. First, the outlier detection and imputation algorithms use general-purpose time series methods rather than epidemiologically-informed models. Disease-specific approaches (e.g., accounting for intervention campaigns, climate-driven seasonality) could improve detection accuracy. Second, the logical consistency checks currently only flag violations without automated correction—resolution requires human review. Third, the system is tailored to Uganda’s administrative hierarchy and DHIS2 configuration; adaptation to other countries would require regeneration of the metadata tables. Fourth, real-time validation of the imputation approach against ground-truth facility records has not been performed, as such records are rarely available. Fifth, the outbreak detection application uses relatively simple kernel-smoothing methods that could be enhanced with more sophisticated approaches to seasonal adjustment and trend detection.

### Broader applicability

Although developed for malaria in Uganda, the architectural patterns—metadata-driven extraction, automated cleaning with provenance tracking, versioned storage, containerized deployment—are applicable to any disease surveillance system built on DHIS2 or similar platforms. The separation of the metadata package from the ETL logic means that adapting the system to a new country or disease domain primarily requires regenerating the metadata tables rather than rewriting pipeline code. The indicator crosswalk pattern is particularly relevant as countries continue to migrate between DHIS2 versions.

### Impact on malaria program operations

Since deployment, the system has transformed several aspects of NMED’s analytical workflow. Analysts access a standardized, cleaned dataset rather than performing independent data processing, eliminating methodological inconsistencies across reports. The outbreak detection dashboard provides weekly situational awareness that was previously unavailable. The version-controlled, reproducible nature of the pipeline has increased confidence in the data underlying policy recommendations. The system has supported the development of routine reports, district-level stratification analyses, and rapid assessment of commodity stock-outs and treatment coverage.

## Conclusion

We have presented an open-source ETL system and metadata package for processing routine health facility malaria data from DHIS2 in Uganda. The system addresses the practical challenges of data extraction, quality assurance, versioning, and deployment that stand between raw facility reports and actionable surveillance intelligence. By making the complete system—including code, metadata, deployment scripts, and a proof-of-principle analytical application—openly available, we aim to provide both a functional tool for Uganda’s malaria program and a reusable template for similar efforts in other settings. The sustained use of this system by Uganda’s NMED demonstrates that systematic, automated data processing can enable responsive, evidence-based program management.

## Data Availability

All data produced in the present work are contained in the manuscript

https://github.com/dd-harp/uga-etl-facility-data

## Acknowledgments

This work was developed through the Robust Analytics for Malaria Policy (RAMP) collaboration between the University of Washington, Uganda’s Department of Health Information, and the National Malaria Elimination Division. We thank the DHIS2 team and the broader open-source health informatics community for the tools upon which this system is built.

## Notes

### Competing Interest Statement

The authors have declared no competing interest.

### Author Declarations

Uganda Department of Health Information waived ethical approval for this work

### Funding

- Gates Foundation: INV 030600

